# Leveraging Health Information System Maturity Assessments to Guide Strategic Priorities: Perspectives from African Leaders

**DOI:** 10.1101/2024.02.27.24303453

**Authors:** Phiona Vumbugwa, Nancy Puttkammer, Moira Majaha, Sonora Stampfly, Paul Biondich, Jennifer Ellen Shivers, Kendi Mburu, Olusegun O. Soge, Chris Longenecker, Jan Flowers, Caryl Feldacker

## Abstract

**Introduction:** Central to a functional public health system is a strong health information ecosystem and robust data use. Many low-and-middle-income countries (LMICs) face the task of digitizing their health information systems (HIS). For health leaders, deciding what to prioritize when investing in HIS strengthening is central to this daunting challenge.

**Objectives:** The study explores how HIS maturity assessment contributes to HIS strengthening, describes the facilitators and barriers to HIS maturity assessments, and how health leaders can prioritize conducting maturity assessments.

**Methods:** This descriptive qualitative study employed key informant interviews (KIIs) with fourteen eHealth leaders at national and international levels working or supporting Ministries of Health’s national HIS in LMICs. Results were analyzed using Dedoose Version 9.0 to develop themes based on the health systems’ building blocks as a framework for identifying facilitators and barriers to conducting HIS maturity assessment.

**Results:** Participants identified maturity assessments as a critical beginning step to HIS strengthening, showing the system’s performance, and building a baseline response to systematic data quality challenges. Barriers to conducting HIS maturity assessment include lacking collaborators’ buy-in, fragmented vision, low financial/human resources, and overdependence on donor priorities. Non- supportive policies, a lack of execution champions, and an inadequately skilled workforce in conducting maturity assessments or negotiating for their prioritization hinder maturity assessment implementation. Frequently identified facilitators to promoting HIS maturity assessment include multi-stakeholder engagement, understanding the country’s HIS ecosystem, and priorities to appropriately integrate maturity assessment objectives. Recommendations include capacity building in data use and conducting maturity assessments at all health system levels to grow the demand and value of HIS maturity assessments.

**Conclusion:** Promoting HIS maturity assessments can help leaders prioritize areas to improve in the HIS ecosystem, making appropriate decisions that steward HIS maturity advancement. Addressing challenges that hinder HIS assessment implementation holds promise to identify a pathway to a strengthened health system.

**Author Summary:** Our manuscript specifically spotlights the perspectives of African eHealth leaders, centering voices on the barriers and facilitators to planning and implementing HIS maturity assessments. We demonstrate their perspective on how conducting maturity assessments can inform understanding of gaps to address in the HIS and strategic direction. We detail the leaders’ recommendations for using HIS maturity assessments in strengthening HIS governance and overall health systems for better population health outcomes in LMIC settings.

## INTRODUCTION

Public health information systems are critical for health systems strengthening (HSS). Global health organizations endeavor to create digitized and integrated health information systems (HIS) with the capacity to collect, collate, and analyze vast amounts of information for rapid response to public health needs (1–3). The COVID-19 pandemic exposed the gaps in HIS’ ability to share information for decision-making (4). The lack of interoperable information systems and the limited implementation of automatic data exchange threaten health systems’ functioning and performance.

Digital HIS provides fast, reliable, and efficient ways for governments to track public health interventions. Several low-and-middle-income countries (LMICs) developed national digital health strategies utilizing the WHO-International Telecommunications Union (ITU) National eHealth Strategy Toolkit, with strategic objectives for developing a more informatics-savvy health organization (ISHO) (5–7). An informatics-savvy health organization obtains, effectively uses, and securely exchanges information to improve public health practice and population health outcomes (8,9).

Strengthening HIS and achieving ISHO goals requires understanding strengths, gaps, and maturity supported by an appropriate framework to effectively track and assess the core functions and capabilities of an HIS (10,11). HIS maturity assessments are often conducted as part of a governance function to learn and provide evidence for decision-making. We define maturity as the degree to which a digitized HIS is interoperable, scalable, offers security and privacy, complies with healthcare standard regulations, and makes health information readily available (4,8). Assessing the digital health systems’ maturity level is important to know what has been tried and/or done to scale or what still needs to be achieved as part of the strategic objectives. Public health leaders need proven tools to assess the maturity of their HIS.

Despite these assessments playing an essential role in HIS strengthening, the facilitators and barriers to conducting maturity assessments are unknown. Conducting maturity assessment supports the implementation of the WHO-ITU National eHealth Strategy toolkit, which shows promises to make HIS improvement plans (12). Few countries (Kenya, Ethiopia, Ghana, and Zambia) have conducted maturity assessments; these have not been conducted consistently and with no follow-up to verify that recommendations were implemented (13–15). Understanding how to plan and implement HIS maturity assessments is a critical step to holistic HSS.

The health system’s functioning is aligned with the six building blocks: governance and leadership, health information, health workforce, financing, medicines and technologies, and service delivery (16). A well-functioning HIS provides information needed for governance and management of health systems, services provision, planning, decision-making, monitoring and evaluation (M&E), and quality improvement of health services (17). We explore (I) health leaders’ perceptions of the value and importance of maturity assessments as part of HIS governance and strategic planning, (II) barriers and facilitators to planning a maturity assessment, and (III) health leaders’ recommendations for overcoming barriers to HIS maturity assessments.

## Methods

### Design

The project used a descriptive qualitative design to assess health leaders’ perceptions of barriers, facilitators, and recommendations for conducting maturity assessments. Key informant interviews were conducted using a structured key informant guide.

### Project setting

The project was conducted with health directors from LMICs participating or supporting a global health informatics leaders’ network. In 2023, I-TECH Digital Initiatives Group (DIGI), in partnership with Regenstrief Institute, launched the eHealth Leaders Forum community of practice (eHLF CoP) for national health information leaders in MoH. The eHLF provides peer learning, networking, and a place to share best practices. Health leaders discuss HIS implementation share challenges faced/opportunities for resources or research, offers peer support in digital/health information systems assessments, planning, and improvement. Through the forum, health leaders expressed the need for support in analyzing and selecting interventions that strengthen HIS.

eHLF is one of several initiatives for HIS capacity building supported through the US Centers for Disease Control and Prevention’s (CDC) Technical Assistance Platform (TAP). Formation and secretariat services for eHLF were supported through TAP. eHLF is part of the overall TAP capacity development strategies that included digital health training for senior and mid-level leaders and the use of informatics-savvy health organization (ISHO) maturity assessments at national and sub-national levels. All MOH respondents participated in eHLF, and some but not all the respondents were exposed to other TAP capacity development interventions.

#### Participant selection, recruitment, and eligibility

Health leaders were selected using a convenience sample from 10 countries. Participants either had a leadership position in the MoH at the director level (n=10) or represented partner organizations funding or supporting digital health innovations in the countries (n=4). The leaders participated in or supported the eHLF.

All participants had at least two years of experience in their roles and thus were expected to be conversant with the health informatics systems or digital health ecosystem. Participants with less than a year of experience in their role and those who did not respond to a second follow-up email were excluded.

An introductory message was developed, and initial contact with countries’ MoH HIS leaders was made through email. where leaders provided their contact numbers to be added to the WhatsApp group platform. After six months of engagement and participation on the forum, at least one health leader per organization and country was purposively selected to participate in a thirty-minute virtual (ZOOM) interview scheduled at the participant’s preferred time.

#### Data collection method

All participants received an initial message seeking consent to participate in the study. After consenting, participants received the KI guide before the interview. A discussion format was used to solicit responses, with participants providing supporting documents where applicable. The interviews were recorded, where 30 minutes exceeded, permission to proceed was sought. Participants answered questions based on their knowledge and shared strategic documents published or grey literature supporting their responses.

#### Data analysis

The interview transcripts were analyzed using Dedoose Version 9.0. Initially, we developed a code book and coding linked to the interview questions. Inductive and deductive themes emerged as we analyzed the codes for each transcript. Inter-coder reliability was performed with primary and secondary coders by defining the codes, testing coding together, independent coding, and discussion after coding. Reliability was tested using Cohen’s kappa formula and coders’ agreement 0.81 of the coding decision. Qualitative thematic analysis was conducted to identify themes relevant to each specific pillar, while content analysis was used to summarize information provided and evidence of best practices to support narratives (18,19). Themes for HIS maturity assessments contribution were matched with best practices drawn from supporting documents. Recommendations on prioritizing and conducting HIS maturity assessment were assigned priority levels from highest to lowest based on the frequency with which the theme was mentioned across respondents.

#### Positionality statement

The lead analyst (PV) is a healthcare practitioner from an LMIC, and their professional experiences, knowledge, and use of health information systems shaped this research. The research was conducted ethically, respecting the perspectives of all participants, contributing to a more inclusive and equitable workspace for individuals of all gender identities. Participants were engaged in a sensitive and open manner.

#### Ethics considerations

The ethics committee of the University of Washington (UW) internal review board (IRB) reviewed and approved the research under the UW IRB STUDY00018156. A formal verbal consent was obtained from all participants prior to conducting an interview. Participants provided consent to record the discussion, which was manually transcribed.

## RESULTS

A total of 14 interviews were conducted; 12 were males, while 2 were females (Table 1). All participants occupied the deputy director level or above in their respective organizations. In the following sections, we report findings under the three objectives. Two categories are used for quotes: 1: MoH who are MoH at the director level (n=10) and 2: PO who are partner organizations funding or supporting MoH (n=4).

**Table 1:** Participant profile.

| *Institution* | *Total* | *National level* | *International level* |
| --- | --- | --- | --- |
| *Ministry of Health* | 7 | 7 | 0 |
| *Universities* | 2 | 2 | 0 |
| *Implementing partners* | 1 | 1 | 0 |
| *Funding Partner* | 2 | 0 | 2 |
| *Global organization* | 2 | 0 | 2 |
| ***Total*** | **14** | **10** | **4** |

### Objective 1: HIS maturity assessment contribution to strengthening HIS

Maturity assessments were identified as a critical beginning step to strengthening HIS. However, all participants agreed that there is a huge gap in conducting HIS maturity assessments in their countries as a national approach, with only 3 out of 10 countries attesting to having conducted an HIS maturity assessment in the past 3 to 5 years. Similarly, one participant stated that strategic planning for HIS maturity assessments has not been done nor prioritized at the continent level. Some participants confirmed being involved in implementing HIS maturity assessments as a once-off, research-based, or nationwide activity. Respondents cited that most of these initiatives were donor-driven; hence, they lacked follow-up to recommendations, ownership of results, and did not prioritize countries’ specific needs.

Participants indicated that a maturity assessment contributes to knowing the system’s performance and understanding the gaps and strengths to build a baseline on which to strengthen HIS. Other participants acknowledged that maturity assessments provide a platform to respond to systematic challenges and recommendations on data quality seen through DQAs and M&Es, which most organizations support. Table 2 summarizes the contribution of HIS maturity assessments to strengthening HIS and provides examples of corresponding best practices from evidence cited by participants.

**Table 2:**
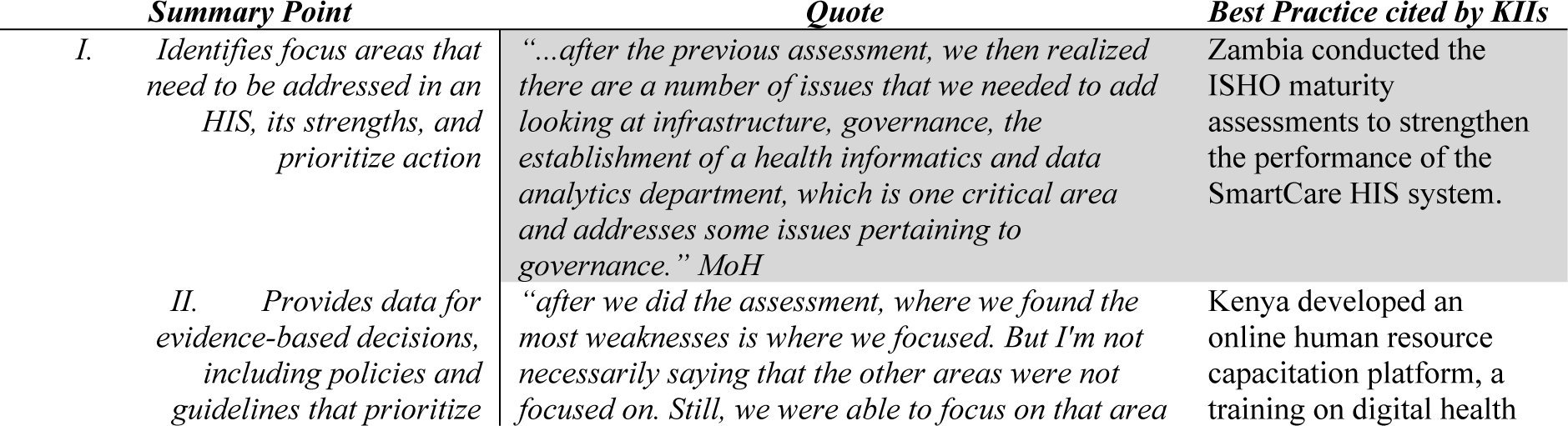
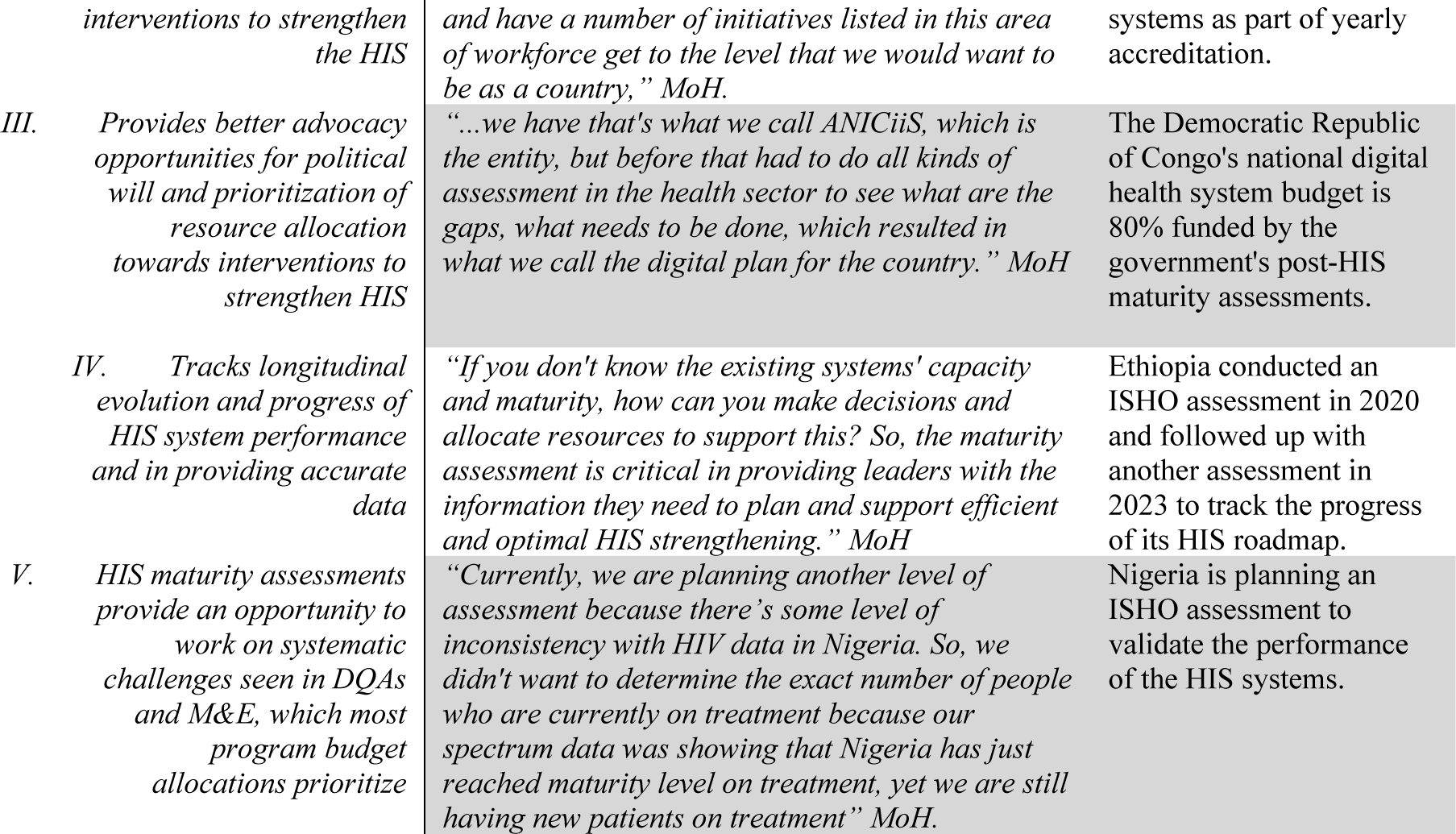
Summary of HIS maturity assessments contribution to HIS strengthening.

**Table 3.** Summary of barriers and facilitators to planning HIS maturity assessments.

| **HSS Pillar** | **Barriers** | **Facilitators** | **Example of HIS strategic documents** |
| --- | --- | --- | --- |
| *Leadership and governance* | Lack of collaborators' buy-in, shared vision, and supporting policies | Coordinated implementation of HIS policies through a national HIS ministry | Ministry of Public Health, DRC. National Development Plan Health Informatics 2020-2024 (20) |
| *Financing* | There is no specific national HIS budget to support HIS maturity assessments. | Allocate a specific national digital/HIS strengthening budget | Democratic Republic of The Congo Ministry of Public Health Quantified Roadmap of Digital Health Investments (21) |
| *Information systems* | Fragmented systems that rely on power and connectivity | Open Source Systems that are interoperable and standardized building from external collaborations | Ministry of Public Health, DRC. National Development Plan Health Informatics 2020-2024 (20) |
| *Workforce* | Inadequate technically capacitated staff to properly understand, use, maintain, and manage HIS infrastructure and technology | Curriculum for HIS workforce and health workers orientation | Kenya's Virtual Academy online training for health workers (22) Zimbabwe and Ethiopia informatics training within the University of Zimbabwe and the University of Gondar |
| *Service delivery* | Competing priorities | Collaborating with other organizations and agencies, | DRC's One Health HIS system |
|  | Siloed and fragmented HIS systems divorced from the national HIS structure make choosing or integrating systems to focus maturity assessment difficult. | leveraging partnerships to provide resources, expertise, and support for HIS strengthening efforts. | Zambia's SmartCare System |
| Medical supplies | lack of standard/user-friendly tools<br>Costly equipment and infrastructure, including maintaining server rooms, providing reliable connectivity at all levels | Flexible resource allocation systems | Integration of mobile technology to supplement HIS |

**Table 4:** Summary of recommendations of ways health leaders can better plan for HIS maturity assessments.

| *Recommendations when planning a maturity assessment* | *Key quote* | *Priority* |
| --- | --- | --- |
| *Collaborative engagement through stakeholder mapping* | *"A multi-stakeholder, a multi-organization, multidisciplinary approach to the assessment, so ensuring that there's a lot of good participation from a key set of diverse stakeholders..." PO* | High |
| *Identify the country's priorities and choose appropriate implementation tools, implementers, time, and process.* | *"And finally, relevance to the local context, so, for the assessment to work well, it should be relevant to that local context so that findings apply to those specific needs of each country or organization. The data collection tool itself should be flexible to meet those needs...." MoH* | High |
| *Increase utilization of available health data to grow the demand and value of an HIS and have its assessment prioritized.* | *"I think we need to be very specific in the way that we solve the problems by using tools, for instance, these same digital health maturity tools, break them down even to the lowest level and keep ourselves alive all the time, like the way that people report monthly, or weekly, to keep health information systems alive or used through reporting and assessments...." PO* | High |
| *Capacity building at all levels to ensure scalability and continuity of the assessment process* | *"One is that country leaders need capacity building and more sensitization. Some of these health leaders don't understand some of this terminology, and the leaders need to be sensitized on the Health Information Systems used for them to give more support." MoH* | High |
| *Identify DQA and M&E planned and include an HIS assessment to provide a cost-effective approach to conducting assessments that can be implemented at routine intervals.* | *"...also, being flexible in the data collection and in the assessment tool itself and applying scalability to already existing plans like M&E and DQA conducted by partners can be cost-effective and promote continuity of HIS maturity assessment, to address different types of systems sizes and complexities and having a cost-effective approach to conducting assessments..." MoH* | Medium |
| *Advocate for increased political will, government ownership, inclusion in strategic plans, and investment into HIS maturity assessments* | *"I feel like the best recommendation I would have is having the political will for and understanding the need for accurate data, then it is easy to mobilize for resources to ensure that maturity assessments are planned and happening because there is some form of accountability" MoH.* | Low |

### Objective 2: Facilitators and barriers to HIS maturity assessments Governance and Leadership

Government support and political will were identified as the main facilitators to promote HIS maturity assessment. Policies and best practices backed by the MoH’s HIS priorities were noted as effective motivators for HIS maturity assessment implementation. Policies reportedly empower health leaders by creating an enabling environment and allowing resource allocation to build a strong base for assessing HIS.

*“At the national level, a policy could create a national health information exchange between agencies, public and private, and would help to improve the coordination of tracking implementation through HIS maturity assessment.” PO*

Participants cited the lack of collaborators’ buy-in and lack of shared vision as barriers to HIS maturity assessments. Without partner support, strategic plans are not enough to encourage engagement in HIS assessments. One respondent stated that,

*“Once we have that five-year strategic plan, we pick out key flagship activities, and we got this digitalization and the strengthening of the health information systems assessments. Once we do that, we engage with the member states ministries to get their buy-in, but you will not probably get 100% of the support, which is like one weakness or challenge because we don’t impose ourselves.” MoH*

Another respondent echoed the lack of support from the government and policymakers and its impact on HIS assessments and strengthening.

*“In some countries, there is a lack of shared vision to invest in HIS. This can make it difficult to get the resources needed to plan maturity assessment and sustain HIS strengthening.” MoH*

#### Health Financing

The availability of dedicated funding for HIS was seen as a conduit to facilitate HIS maturity assessments. Participants acknowledged the advocacy surrounding the importance of HIS and its benefits, with some governments beginning to fund HIS interventions and infrastructure. Three participants from national and international levels gave an example of best practice in the Democratic Republic of Congo, where the government finances 80% of digital health information systems, with 1.5% of the national health budget invested towards strengthening digital health systems.

All participants firmly acknowledged donor support, including the development of district health initiatives and electronic health records (EHR) systems, as important to start conversations around maturity assessment and assessing scalability. One participant stated,

*“The openness by the government to receive financing from donors for the assessments, that’s an opportunity…” MoH*

All participants acknowledged that financing for HIS remains the most significant challenge impacting efforts to prioritize planning of HIS assessments and overall system strengthening. It was commonly reported that a specific national HIS allocated budget was lacking. One key informant stated,

*“Looking at advocacy around M&E has been shown partly by having these critical positions in place, but is not complimented by a budget to say if we have at least 8-10% of the Ministry budget committed to M&E and informatics, then we know that the oil of the system is guaranteed. So, we find that advocacy is not there in terms of its translation into tangible activities due to poor funding. So, I would advocate that 8-10% of the Ministry budget always support the HIS’s M&E, including assessments, health informatics, and systems strengthening.” PO*

Most participants agreed that over-reliance on donors and funders prevents flexibility in planning for HIS assessments, which are generally not planned as part of the restricted funding. The same is seen to have placed over-dependency on donor priorities with less prioritization or room to negotiate for financing other critical competing priorities like maturity assessments. In support, a few participants indicated that their country’s budget to roll out the District Health Information System (DHIS2) is funded by the Global Fund and PEPFAR. One respondent stated,

*“In terms of specific budget to support, like DHIS2 hardware and infrastructure, that has been donor funded. Much of the support for DHIS2, if not mistaken, about 80% comes from the Global Fund, 20% is from PEPFAR, and 0% is from the government. Right now, we are rolling out EHR, electronic medical record; much of the support is coming from PEPFAR followed by Global Fund, and assessing their maturity is not part of the grant” KII 11 (MoH)*

Several participants indicated that having outside funders as the leading financial supporters severely impacts the ability of a country to prioritize and promote interventions that support HIS maturity assessments, as donors dictate the priority of the funding. One participant agreed, saying,

“*…what is not going well is that the financing structure has been too donor oriented, so the priorities have been donor orientated*” *MoH*.

#### Workforce

Participants noted the lack of training and skillset in maturity assessments for health workers who are essential health data collectors and users as a barrier to collecting valuable and credible data for prioritizing HIS assessment planning. One participant supported saying,

*“…skills and training of health workers in maturity assessment are limited, and that is a barrier, as it means that they may not be able to provide the most up-to-date information to show the performance of the system; there is a delay in getting this data to make decisions, delaying planning on appropriate HIS interventions” MoH*.

Skilled informatics and maturity assessment workforce were cited as vital in successfully implementing HIS assessments, yet countries lack personnel who can lead or conduct HIS assessments. Most health directors cited a lack of recognition for health informatics roles within the health workforce and a lack of power to negotiate priorities to focus on HIS being a barrier to HIS assessments. The lack of motivation and poor working conditions, including long hours, low pay, and inadequate resources, contributed to governments’ inability to retain key informatics skilled staff.

It was commonly noted that countries lack specific training structures for health informatics or HIS personnel as digitization of HIS has only recently become part of the health system structure. Two participants from national universities echoed that training has often been ad-hoc, and developing standardized training programs is challenging. Respondents believed that improving these conditions could motivate leaders to invest in the HIS workforce, which is key in HIS assessments and overall healthcare improvement.

Most participants, 11 of 14, echoed that the current workforce structure facilitates the capacity of available health workers with knowledge of the importance of conducting HIS maturity assessments. Participants cited methods such as including HIS mentorship or training in license renewal platforms, departmental mentorships to review reports, data use, and structuring tailored training on HIS for specific needs. All the above were seen as facilitators to appropriately integrate maturity assessment objectives as the workforce understands the country’s HIS ecosystem and priorities better, thus making planning easy. Across the board, all respondents noted that the workforce’s skillset is essential in ensuring a well-functioning HIS with the ability to plan and conduct assessments effectively, implementing any required changes noted.

#### Infrastructure and Medical Supplies

Most participants believed the diversity of systems, the heterogeneity in investments in ICT infrastructure, and HIS supplies to be significant barriers for many LMICs to plan for HIS maturity assessments due to cost and availability. Also, the expensive technology has led to having several fragmented HIS systems, either disease-specific or program-specific and non-interoperable; as such, integrating HIS maturity assessment plans becomes a challenge. Many cited this as why HIS is not prioritized over other health system pillars. The HIS infrastructure should be defined to plan for a maturity assessment, as explained by one participant,

*“I think the infrastructure is a real challenge because we need to ensure that we have connectivity, which is one of the challenges, apart from the equipment like servers, firewalls, and switches that are too expensive to buy and maintain, thus makes HIS assessment less priority, we only replace the piece not functional” MoH*.

A few participants brought up an important point that for the few countries that have made steady progress in planning and implementing HIS assessments, the biggest challenge has been having standard/user-friendly tools, standardizing and having interoperable systems as countries use different electronic medical records (EMR) systems, such as open source medical records system (OpenMRS), laboratory and management information systems (LAMIS), and others. Many participants stated that each implementing partner would have its own unique system, which has resulted in a very disintegrated HIS landscape, making it challenging to prioritize HIS maturity assessment planning as systems need to be separated because their maturity and implementation may not be comparable.

*“So, there are multiple systems that are in use and coming, and they are not interoperable. As a result, there is duplication and redundancy in that aspect; we find it hard to pick which one to strengthen.” PO*

Uniquely, one participant pointed out that infrastructure for health information systems falls under different departments and is regulated by ministries, like the Ministry of ICT or Finance, making it challenging for the MoH to prioritize HIS funding and planning HIS maturity assessments as the infrastructure belongs to a different ministry. Stating something almost similar, several respondents cited technology evolving quickly and infrastructure becoming outdated or incompatible faster than strengthening processes can catch up, posing a challenge for keeping HIS assessment planning and implementation up to speed.

### Objective 3: Ways health leaders can better plan for HIS maturity assessments

Participants provided various ways health leaders can better plan HIS maturity assessments in their countries.

All participants recommended multi-stakeholder collaborative engagement when planning maturity assessments, from the idea’s conception to completion. Several participants attested that engagement is crucial to ensure that all parties understand and agree on the objectives and methodology of the assessment, leverage existing M&E systems, and piggyback on already established systems, such as the DHIS2 monitoring, to effectively plan for an HIS maturity assessment. One participant emphasized that for HIS maturity assessment planning to be effective, there needs to be strong collaborative engagement of all stakeholders to advocate for government prioritization and political will to support the initiative.

Several participants stated that collaborators should be involved in planning a maturity assessment based on their context. Another participant stated that stakeholders hold different powers and expertise, which is key when planning a maturity assessment for the country. Participants outlined the need to bring a sense of recognition, ownership, and support to plan a participatory action-based maturity assessment. More than half of the participants supported planning for action by prioritizing using context-based assessment tools, minimizing duplication of activities, and letting countries decide the processes. In support, one participant echoed that.

*“Stakeholders should not be restricted only to MoH’s HIS and digital health department but include all partners, implementing, funding, Ministry of Information and the regulatory bodies, telecommunications (public and private), power supply organizations, community representatives and advocates. The inclusion of such key stakeholders capacitates them to understand the need for prioritizing HIS assessments and their role in setting goals and ownership of recommendations.” MoH*

Almost all (11/14) participants highly prioritized increasing the use of data when planning for HIS maturity assessment. Respondents indicated that the available systems’ data should be used to show the system’s weaknesses or strengths for the leaders to focus the assessment. A few also cited that stakeholders should have access and the ability to analyze or report the data, and that would prepare them to understand assessment findings and take ownership to improve the system. Table 2 summarizes recurring recommendations from participants.

## DISCUSSION

While HIS leaders recognized the value of having evidence from HIS maturity assessments to guide them in planning for HIS strengthening, most felt significant barriers to conducting such assessments. To achieve a functional, optimized, sustained, and strengthened health system, HIS maturity assessments provide a critical beginning step to a system’s performance status, highlighting areas to integrate, expand, and scale up. Key facilitators to implementing HIS maturity assessment included coordination, collaborating with existing M&E programs, and knowledgeable health workers to conduct HIS assessments at all health facilities. Barriers to implementing maturity assessments include a lack of skilled workforce knowledgeable in HIS maturity assessments, fragmented HIS systems using expensive infrastructure, and lack of financing. Addressing these barriers and facilitators is crucial for achieving effective HIS strengthening and data-driven decision-making in healthcare systems.

Governance is critical to HSS; maturity assessments, especially participatory assessments, can help strengthen that. Weak health systems governance in LMICs has resulted in fragmented or ad hoc health policy formulation, poising challenges in implementing HIS maturity assessments and impacting efforts to strengthen overall health systems (23,24). For example, leadership and governance for HIS include having an eHealth Technical Working group that oversees the implementation of digital health, interoperability activities, and financial resourcing to aid the implementation of recommendations from the assessment (15,25). Information derived from maturity assessments can benefit HIS governance in (a) identifying issues, b) providing guidance for improvement in health systems’ policies, and (c) improving efficiency, effectiveness, performance, and productivity in the whole health system (2). Through participatory planning, health systems governance leadership in Ghana and Rwanda effectively prioritized areas to improve in their HIS, supported by strong governance structures (26–28). There is a need for policies supporting HIS maturity to strengthen systems. For a health system to function optimally, continuous monitoring and evaluation of the system is required.

There is a need to grow the knowledge about the value of an HIS maturity assessment. Making maturity assessments routine and operationalized as part of a strategic vision could increase demand for sustainable HIS assessments. Efforts should focus on increasing the need for data use and efficient health systems, thus building the culture of conducting HIS maturity assessments using integrated and decentralized approaches. First, strengthening the capacity of health leadership in planning and conducting systems performance monitoring at all health facilities promotes accountability to health data, increasing data use. When the health team understands their responsibility and accountability, teamwork is cultivated, which is essential to improving data use, quality for informed decision-making, policy change, and planning (4,26,29). Second, having multisectoral HIS steering committees, developing HIS interoperability roadmaps, and creating a costed work plan could strongly generate the demand for HIS maturity assessments (30). Participatory planning addresses not only technical aspects but also the cultural, structural, and governance-related factors to having an effective maturity assessment. Third, when planning HIS maturity assessment, collaborative efforts should leverage existing M&E systems or services to co-develop the goals/objectives of the assessment based on country needs, priorities, and collaborators’ implementation efforts (31). The benefits and value of conducting HIS maturity assessments are realized when the country translates the recommendations into binding policies and HSS activities.

HIS maturity assessments are critical to establishing an evidence base and process for systematically prioritizing objectives in the health sector. Health programs focus resources, de- duplicate work, and reduce staff workload, potentially strengthening health systems. Because the HIS landscape and context will evolve over time, assessments should not be conducted as a one-time marker but as part of a routine iterative cycle for understanding the HIS, feeding into updates to the strategic vision, strategic objectives, and action plans for maturity. To achieve this vision of sustainable HIS assessments, it is imperative that leaders have a shared vision and skilled champions to plan/implement the activities, financing, and coordination. Investments in health have been donor-driven and fragmented, particularly in information systems in sub-Saharan African countries, which has resulted in a lack of shared vision and drivers for HIS assessments (32,33). To overcome this barrier, sustainable HIS maturity assessments require strong buy-in and leadership from governments, with sufficient consultations among key stakeholders to support better planning and implementation of maturity assessments (34,35). This approach can be a pathway to ensuring the results will be relevant and useful to all critical partners supporting HIS beyond donor-driven investments and projects.

## Limitations

Most countries had not conducted an HIS maturity assessment at the time of interviews, so participant knowledge was based on M&E or demographic health surveys, which did not focus on HIS. Secondly, most (80%) key informants were from Africa, and all were engaged through eHLF, so they may not have represented all health system leaders in LMICs. However, we expect the barriers, facilitators, and recommendations they named would resonate with other LMIC regions. Thirdly, the research did not ask about the drawbacks of conducting maturity assessments or why they do not bring value to HIS, strengthening the assumption that health leaders think HIS maturity assessments are important. Lastly, the structure of questions resulted in confounding responses, with some participants treating HIS strengthening and HIS assessments interchangeably.

## Conclusion

Strengthening health information systems is vital in improving healthcare for all in LMICs. With the growing access to technology and increasing demand for digital health solutions, assessing the maturity of HIS to aid in identifying digital health priorities plays a vital role in improving HSS. Countries still face challenges in conducting HIS maturity assessments and operationalizing results to strengthen their HIS. The challenges include lack of prioritization of HIS due to low political will, a lack of shared vision due to the donor-dependent funding of HIS, and a lack of essential skills in the health workforce to conduct maturity assessments. Addressing these barriers is crucial for planning for and executing HIS maturity assessments, potentially achieving effective HIS strengthening through data-driven decision-making in healthcare systems. Key to planning an effective HIS maturity assessment includes multi-collaborative engagements, contextualizing to country needs/priorities, using existing resources/structures or M&E plans, advocating for government prioritization, and gaining political will. Institutionalizing HIS maturity assessments as part of HIS governance offers a promise to adopt and build a foundation for having interoperable, integrated, and sustainable HIS integral to a well-functioning and strengthened health system.

## Declarations

### Competing interest statement

The authors have declared that no competing interests exist.

### Financial disclosure statement

The study did not receive any funding.

### Data Sharing

All data to the study is stored in a secure folder in SharePoint drive under the University of Washington. Data is only accessible through a request to the study team. This data can be made available to reviewers upon request. A link to the dashboard may be made available to authorized reviewers as it contains patient information that cannot be shared widely according to patient rights and confidentiality of information regarding health data.

## Data Availability

All data to the study is stored in a secure folder in SharePoint drive under the University of Washington. This data can be made available to reviewers upon request. A link to the dashboard may be made available to authorized reviewers as it contains patient information that cannot be shared widely according to patient rights and confidentiality of information regarding health data.

## Acknowledgments

The authors would like to extend their gratitude to all the HIS leaders for their invaluable contributions instrumental to accomplishing the study objectives. Second, we acknowledge the support from PATH and the US Centre for Disease Control (US CDC) to the Technical Assistance Partnership (TAP), which provided support for launching the eHealth Leaders Forum (EHLF). Lastly, sincere gratitude to the research team who contributed to the successful completion of this study.

## Notes

### Competing Interest Statement

The authors have declared no competing interest.

### Funding Statement

The author(s) received no specific funding for this work.

### Author Declarations

The ethics committee of the University of Washington (UW) internal review board (IRB) reviewed and approved the research under the UW IRB STUDY00018156

